# Antihypertensive Medication Class and Incident Alzheimer’s Disease and Related Dementias: External Replication Across Two Independent Healthcare Data Sources-the Vanderbilt EHR Synthetic Derivative and US Medicare Claims

**DOI:** 10.64898/2026.09.11.26362859

**Authors:** Anshul Tiwari, Rui Chen, Yuting Tan, Yan Yan, Zhexing Wen, Wei-Qi Wei, Xue Zhong, Bingshan Li

## Abstract

**Importance:** Hypertension is among the most important modifiable risk factors for Alzheimer’s disease and related dementias (ADRD), but whether different antihypertensive drug classes confer different cognitive risk, particularly through differential modulation of angiotensin II receptor signaling, remains uncertain. Prior pharmacoepidemiologic evidence is based largely on a single Medicare cohort, and external validation across distinct healthcare data environments is needed.

**Objective:** To externally replicate and extend prior findings across two structurally independent healthcare data sources, evaluating whether predominant use of antihypertensive medications that stimulate versus inhibit angiotensin II type 2 and type 4 receptor signaling is associated with risk of incident ADRD.

**Design, Setting, and Participants:** Retrospective new-user, active-comparator cohort study with propensity-score matching, conducted in parallel across two independent data sources: the Vanderbilt University Medical Center Synthetic Derivative (VUMC SD; deidentified academic medical center electronic health record through February 2025) and the Merative^™^ MarketScan^®^ Medicare Database (2015–2023). Adults aged 65 years or older with hypertension and at least 365 cumulative days of antihypertensive exposure were included after a 365-day blanking period.

**Exposures:** Predominant use (≥80% of cumulative exposure days) of receptor-stimulating antihypertensive medications (angiotensin II receptor blockers, dihydropyridine calcium channel blockers, thiazide diuretics) vs receptor-inhibiting antihypertensive medications (angiotensin-converting enzyme inhibitors, β-blockers, nondihydropyridine calcium channel blockers); a third group of nonusers comprised participants without predominant use of either class.

**Main Outcomes and Measures:** Time to first incident ADRD diagnosis was identified using ICD-9 and ICD-10 codes consistent with the CMS Chronic Conditions Data Warehouse definition. Adjusted hazard ratios (HRs) and 95% CIs were estimated using Cox proportional hazards models with time-dependent covariates.

**Results:** The Medicare cohort included 1,596,034 beneficiaries (mean [SD] age, 75.0 [8.0] years; 55% female), and the VUMC SD cohort included 80,267 patients (mean [SD] age, 72.3 [5.8] years; 57% female). Over approximately 7 years of follow-up, ADRD incidence was lower among predominant users of stimulating vs. inhibiting medications in both cohorts (Medicare: 6.7% vs. 8.2%; VUMC SD: 4.9% vs. 5.7%). After adjustment, predominant use of stimulating medications was associated with lower ADRD risk versus inhibiting medications in both cohorts (Medicare: HR, 0.92; 95% CI, 0.90–0.93; *P* < .001; VUMC SD: HR, 0.92; 95% CI, 0.85–0.99; *P* = .03). Established cardiovascular and neuropsychiatric risk factors (atrial fibrillation, heart failure, chronic kidney disease, depression) showed expected associations with ADRD risk in both cohorts, supporting internal validity.

**Conclusions and Relevance:** Across two structurally independent data sources, the predominant use of antihypertensive medications that stimulate angiotensin II type 2 and type 4 receptor signaling was associated with a modestly lower risk of incident ADRD compared with predominantly inhibiting agents. By replicating in two distinct data environments, these findings extend prior single-source pharmacoepidemiologic evidence and support evaluation of receptor-targeted antihypertensive strategies for cognitive outcomes in randomized comparative-effectiveness trials.

## Introduction

Alzheimer’s disease (AD) is the leading cause of dementia worldwide, affecting more than 6 million adults in the United States, with projections approaching 13 million by 2050. Annual healthcare and caregiving costs exceed $300 billion.^1^ Although recent anti-amyloid-β monoclonal antibodies represent an important therapeutic advance, scalable preventive strategies remain an urgent public health priority. Contemporary models conceptualize AD as a multifactorial neurovascular disorder in which amyloid-β deposition and tau pathology coexist with neuroinflammation, oxidative stress, blood-brain barrier disruption, and chronic cerebral hypoperfusion.^2-7^

Hypertension is the most consistently identified vascular contributor to cognitive impairment and dementia. Midlife hypertension is robustly associated with late-life cognitive decline and AD,^8-11^ and the 2020 Lancet Commission on Dementia Prevention identified hypertension as a key midlife modifiable risk factor. In SPRINT MIND, intensive systolic blood pressure lowering significantly reduced incident mild cognitive impairment and the composite of MCI or probable dementia.^12-14^ Meta-analyses of antihypertensive randomized trials likewise show a modest reduction in incident dementia, although class-specific differences remain debated.^15^

Beyond systemic blood pressure regulation, the renin-angiotensin system (RAS) is an active signaling pathway in the central nervous system that influences cerebral perfusion, oxidative stress, neuroinflammation, and amyloid metabolism.^16^ Angiotensin II signaling through the type 1 receptor (AT_1_R) drives vasoconstriction, reactive oxygen species generation, endothelial dysfunction, microglial activation, and pro-inflammatory cytokine release-mechanisms implicated in AD pathogenesis.^17, 18^ Angiotensin II signaling through the type 2 receptor (AT_2_R) and the angiotensin IV/type 4 receptor (AT_4_R) appears to oppose AT_1_R activity, promoting vasodilation, anti-inflammatory signaling, neuronal survival, synaptic plasticity, and increased cerebral blood flow.^16,19,20^

Different antihypertensive drug classes engage these pathways differently. Angiotensin receptor blockers (ARBs) selectively antagonize AT_1_R while preserving and indirectly enhancing AT_2_R and AT_4_R signaling, because circulating angiotensin II is redirected to unblocked receptors. Dihydropyridine calcium channel blockers and thiazide diuretics do not directly inhibit RAS signaling and likewise preserve type 2 and type 4 receptor activity. Angiotensin-converting enzyme inhibitors (ACEIs), β-blockers (which suppress renin release), and nondihydropyridine calcium channel blockers reduce downstream activation of all angiotensin II receptors. ACE itself participates in amyloid-β degradation, complicating the net neurochemical effect of ACE inhibition.^16^ Centrally and peripherally acting agents within these classes may also differ in blood-brain barrier penetration, contributing to heterogeneity across observational reports.^21^

Pharmacoepidemiologic studies have suggested that ARBs and other RAS-modulating agents may be associated with lower dementia risk than other antihypertensive classes,^22-25^ but estimates have varied across populations and exposure definitions. More recently, Marcum and colleagues introduced an explicit receptor-pathway framework, reclassifying common antihypertensives by their net effect on AT_2_R and AT_4_R signaling and reporting that predominant use of stimulating agents was associated with lower incident AD risk in a single Medicare cohort using an active-comparator new-user design with propensity-score weighting.^26^ Because that analysis was based on a single national claims dataset comprising only a 5% random Medicare sample, the findings have not yet been externally validated in a structurally different data environment. Independent replication is particularly important for claims-based dementia research because residual confounding, outcome misclassification, and dataset-specific coding practices can each bias single-source estimates in ways that may not generalize to other settings.

We therefore conducted a replication and extension study using two structurally independent healthcare data sources in parallel, with three aims: (1) to replicate the receptor-pathway association in an independent EHR-based cohort drawn from the VUMC Synthetic Derivative,^27^ a deidentified academic-medical-center EHR with longitudinal structured data and clinical notes; (2) to extend the analysis to a contemporary US Medicare claims cohort drawn from the MarketScan Medicare Database (2015–2023), complementing the earlier single-Medicare analysis with contemporary data; and (3) to evaluate the consistency of associations across distinct healthcare data environments using harmonized active-comparator new-user designs. To our knowledge, this is the first dual-source external validation of the receptor-pathway antihypertensive framework for incident ADRD.

## Methods Study Design

We performed a retrospective cohort study using two independent data sources and applying a harmonized new-user, active-comparator design with a 365-day exposure-accumulation window and a 365-day blanking period to reduce reverse causation.^28, 29^ The study design and timing of exposure ascertainment, blanking, and outcome follow-up are summarized in Figure 1.

**Figure 1.**
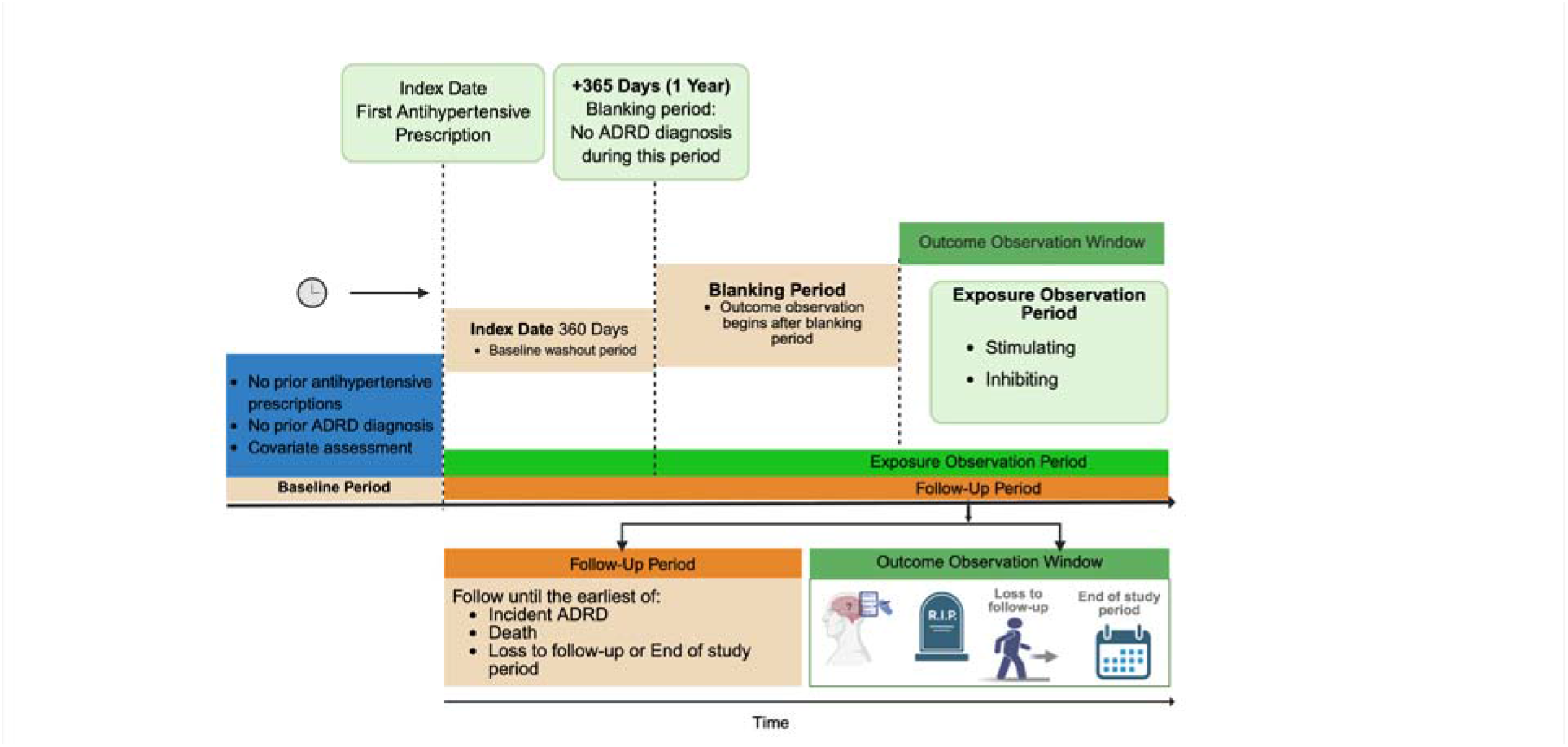
Study design schematic showing baseline period, index date, 365-day blanking period, exposure observation period, and follow-up window.

## Data Sources

Two complementary, structurally independent data sources were used in parallel. By design, the two sources differ in patient populations, geographic coverage, and, critically, in how exposures, comorbidities, and outcomes are ascertained, allowing each cohort to serve as an external check on biases specific to the other.

### VUMC Synthetic Derivative (EHR cohort)

The VUMC Synthetic Derivative is a deidentified mirror of the Vanderbilt University Medical Center electronic health record covering approximately 4 million unique patients and containing structured diagnoses, procedures, encounter histories, laboratory results, medication orders, and free-text clinical notes; all available data through February 2025 were extracted for this analysis.^27^

### Merative MarketScan Medicare Database (2015–2023)

Data for the Medicare analysis were derived from the Merative^™^ MarketScan^®^ Medicare Database for the period 2015–2023. The MarketScan Medicare Database contains deidentified administrative claims data for inpatient, outpatient, and outpatient prescription drug utilization among individuals with Medicare supplemental insurance or Medicare Advantage coverage; for individuals with Medicare supplemental insurance, both Medicare and supplemental insurance paid portions of claims are represented. Study data were extracted using International Classification of Diseases, 9th and 10th Revision, Clinical Modification (ICD-9-CM and ICD-10-CM) diagnosis codes and National Drug Codes (NDC). The MarketScan Medicare Database was selected to enable external replication of the receptor-pathway findings in a contemporaneous, national dataset structurally distinct from the VUMC SD EHR.

### Study Population and Eligibility

Adults with hypertension were identified using ICD-9 and ICD-10 diagnosis codes (eTable 1 in Supplement 1). Eligible participants were required to have (1) at least 12 months of continuous Medicare enrollment or continuous EHR activity before the index date for baseline characterization;at least 365 cumulative exposure days to any qualifying antihypertensive medication; (3) age ≥65 years at the index date; and (4) no prior diagnosis of ADRD before the index date. The index date was defined as the date of the prescription that completed the 365-day cumulative exposure requirement. Continuous treatment episodes were constructed by joining consecutive prescriptions separated by no more than 365 days.

### Exposure Definition

Antihypertensive medications were classified by their net effect on angiotensin II type 2 and type 4 receptor signaling. Receptor-stimulating medications (hereafter, *stimulating*) included ARBs, dihydropyridine calcium channel blockers, and thiazide diuretics. Receptor-inhibiting medications (hereafter, *inhibiting*) included ACEIs, β-blockers, and nondihydropyridine calcium channel blockers. The full list of qualifying medications is provided in eTable 2 in Supplement 1.

Cumulative days of exposure to each receptor-pathway class were computed from prescription fill dates and days-supply. Participants were assigned to one of three time-updated treatment groups: (1) the *stimulating* group, defined as having stimulating medication for ≥80% and inhibiting medication for <80% of cumulative exposure days; (2) the *inhibiting* group, defined as having inhibiting medication for ≥80% and stimulating medication for <80% of cumulative exposure days; and (3) the *nonusers* group, defined as having neither stimulating nor inhibiting medication for ≥80% of cumulative exposure days. The 80% threshold was prespecified to define predominant medication exposure.^30^

### Outcome

The primary outcome was time to first incident diagnosis of Alzheimer’s disease and related dementias. ADRD was identified using ICD-9 and ICD-10 codes consistent with the ADRD definitions used in prior Medicare claims-based dementia studies, requiring at least one qualifying inpatient or outpatient claim (Medicare) or structured diagnosis entry (VUMC SD).^31^

### Covariates

Covariates were selected based on factors associated with both antihypertensive treatment selection and the risk of ADRD. Time-fixed covariates included age at the index date and sex. Additional baseline covariates ascertained at baseline included duration since first documented hypertension diagnosis and indicator variables for atrial fibrillation, ischemic heart disease, chronic heart failure, type 2 diabetes mellitus, obesity, chronic kidney disease, and depression. These conditions were selected based on their established associations with cardiovascular and cerebrovascular outcomes and with risk of cognitive decline, drawing on prior epidemiologic literature. Diagnoses were identified using ICD-9 and ICD-10 codes from outpatient and inpatient encounters (Medicare) or structured EHR records (VUMC SD).

### Statistical Analysis

Baseline characteristics were summarized as means (SDs) or counts and proportions stratified by treatment group. To balance baseline covariates between predominant exposure groups, propensity-score matching was performed within each data source.^32, 33^ Propensity scores for predominant stimulating vs. inhibiting medication use were estimated using multivariable logistic regression that included the baseline sociodemographic characteristics and comorbidities already discussed in the above section. Stimulating, inhibiting, and non-users were matched 1:1 on propensity score.

Time zero for follow-up was set at the end of the 365-day blanking period. Participants were followed until the first occurrence of incident ADRD, death, disenrollment, loss to EHR follow-up, or the end of the study period, whichever came first. Cox proportional hazards regression with time-dependent exposure and confounder updates was used in the matched cohorts to estimate hazard ratios and 95% CIs. The *inhibiting* group served as the reference category for the primary contrast.

## Results

### Cohort Characteristics

After propensity-score matching, baseline characteristics were well balanced across treatment groups in both data sources (Table 1). The Medicare cohort comprised 1,596,034 beneficiaries: 409,682 stimulating users, 409,682 inhibiting users, and 776,670 nonusers. Mean (SD) age was 74.8 (8.04), 74.9 (8.07), and 75.1 (7.80) years, respectively, and the cohort was 55% female. The VUMC SD cohort comprised 80,267 patients: 24,126 stimulating users, 24,126 inhibiting users, and 32,015 nonusers, with mean (SD) age of 72.3 (5.86), 72.4 (5.89), and 72.2 (5.76) years, respectively (57% female).

**Table 1.** Baseline characteristics of patients by treatment group.

| Characteristic | Medicare |  |  | VUMC SD |  |  |
| --- | --- | --- | --- | --- | --- | --- |
|  | Stimulating<br>(N=409,682) | Inhibiting<br>(N=409,682) | Nonusers<br>(N=776,670) | Stimulating<br>(N=24,126) | Inhibiting<br>(N=24,126) | Nonusers<br>(N=32,015) |
| ADRD over 7 y, No. (%) | 27,409 (6.7) | 33,487 (8.2) | 59,526 (7.7) | 1,191 (4.9) | 1,365 (5.7) | 1,743 (5.4) |
| Age, mean (SD), y | 74.8 (8.04) | 74.9 (8.07) | 75.1 (7.80) | 72.3 (5.86) | 72.4 (5.89) | 72.2 (5.76) |
| **Sex, No. (%)** |  |  |  |  |  |  |
| Male | 181,971 (44.4) | 182,963 (44.7) | 350,992 (45.2) | 10,362 (42.9) | 10,250 (42.5) | 13,366 (41.7) |
| Female | 227,711 (55.6) | 226,719 (55.3) | 425,678 (54.8) | 13,764 (57.1) | 13,876 (57.5) | 18,649 (58.3) |
| **Prior medical history, No. (%)** |  |  |  |  |  |  |
| Type 2 diabetes | 28,541 (7.0) | 29,451 (7.2) | 55,269 (7.1) | 1,088 (4.5) | 1,089 (4.5) | 1,453 (4.5) |
| Obesity | 14,191 (3.5) | 14,367 (3.5) | 25,374 (3.3) | 654 (2.7) | 648 (2.7) | 713 (2.2) |
| Chronic kidney disease | 13,728 (3.4) | 15,234 (3.7) | 28,299 (3.6) | 512 (2.1) | 489 (2.0) | 585 (1.8) |
| Depression | 12,472 (3.0) | 15,487 (3.8) | 25,809 (3.3) | 1,125 (4.7) | 1,087 (4.5) | 1,498 (4.7) |
| Ischemic heart disease | 21,043 (5.1) | 21,083 (5.1) | 41,672 (5.4) | 1,824 (7.6) | 1,888 (7.8) | 2,447 (7.6) |
| Atrial fibrillation | 11,849 (2.9) | 11,861 (2.9) | 23,465 (3.0) | 916 (3.8) | 1,041 (4.3) | 1,332 (4.2) |
| Chronic heart failure | 3,750 (0.9) | 3,744 (0.9) | 7,327 (0.9) | 395 (1.6) | 398 (1.6) | 469 (1.5) |

Comorbidity prevalence was similar across treatment groups within each data source (Table 1). In the Medicare cohort, type 2 diabetes prevalence was approximately 7%, obesity 3.3%-3.5%, and chronic kidney disease 3.4%-3.7%. Depression was modestly more frequent among inhibiting users (3.8%) than stimulating users (3.0%) and nonusers (3.3%). Ischemic heart disease (∼5%), atrial fibrillation (∼3%), and chronic heart failure (∼0.9%) were comparably distributed. In the VUMC SD cohort, comorbidity prevalence was generally lower but balanced across groups: type 2 diabetes 4.5%, obesity 2.2%-2.7%, chronic kidney disease 1.8%-2.1%, and depression 4.5%-4.7%; ischemic heart disease (7.6%-7.8%), atrial fibrillation (3.8%-4.3%), and chronic heart failure (1.5%-1.6%) were similarly distributed. The 7-year crude incidence of ADRD was 6.7%, 8.2%, and 7.7% in the Medicare stimulating, inhibiting, and non-user groups, respectively, and 4.9%, 5.7%, and 5.4% in the VUMC SD groups, respectively.

### Discovery and Extension in Medicare

In the Medicare extension cohort, predominant use of stimulating antihypertensive medications was associated with a statistically significant lower risk of ADRD compared with predominant use of inhibiting medications (adjusted HR, 0.92; 95% CI, 0.90-0.93; *P* < .001), consistent in direction and magnitude with prior published estimates.^12^ ADRD risk did not differ meaningfully between nonusers and stimulating users (HR, 1.01; 95% CI, 1.00-1.03; *P* = .14), suggesting the contrast was driven principally by elevated risk among predominant inhibiting users rather than by reduced risk in the stimulating group.

Established cardiovascular and neuropsychiatric covariates showed expected associations with ADRD risk (Figure 2A), supporting internal validity. Atrial fibrillation conferred more than a twofold increased hazard (HR, 2.33; 95% CI, 2.23-2.43); chronic heart failure (HR, 1.65; 95% CI, 1.52-1.79), chronic kidney disease (HR, 1.44; 95% CI, 1.38-1.51), and depression (HR, 1.70; 95% CI, 1.63-1.77) were also strongly associated with ADRD (all *P* < .001). Type 2 diabetes (HR, 1.18), ischemic heart disease (HR, 1.05), and obesity (HR, 1.08) showed more modest associations. Female sex was associated with slightly higher ADRD risk (HR, 1.09; 95% CI, 1.07-1.11; *P* < .001).

**Figure 2.**
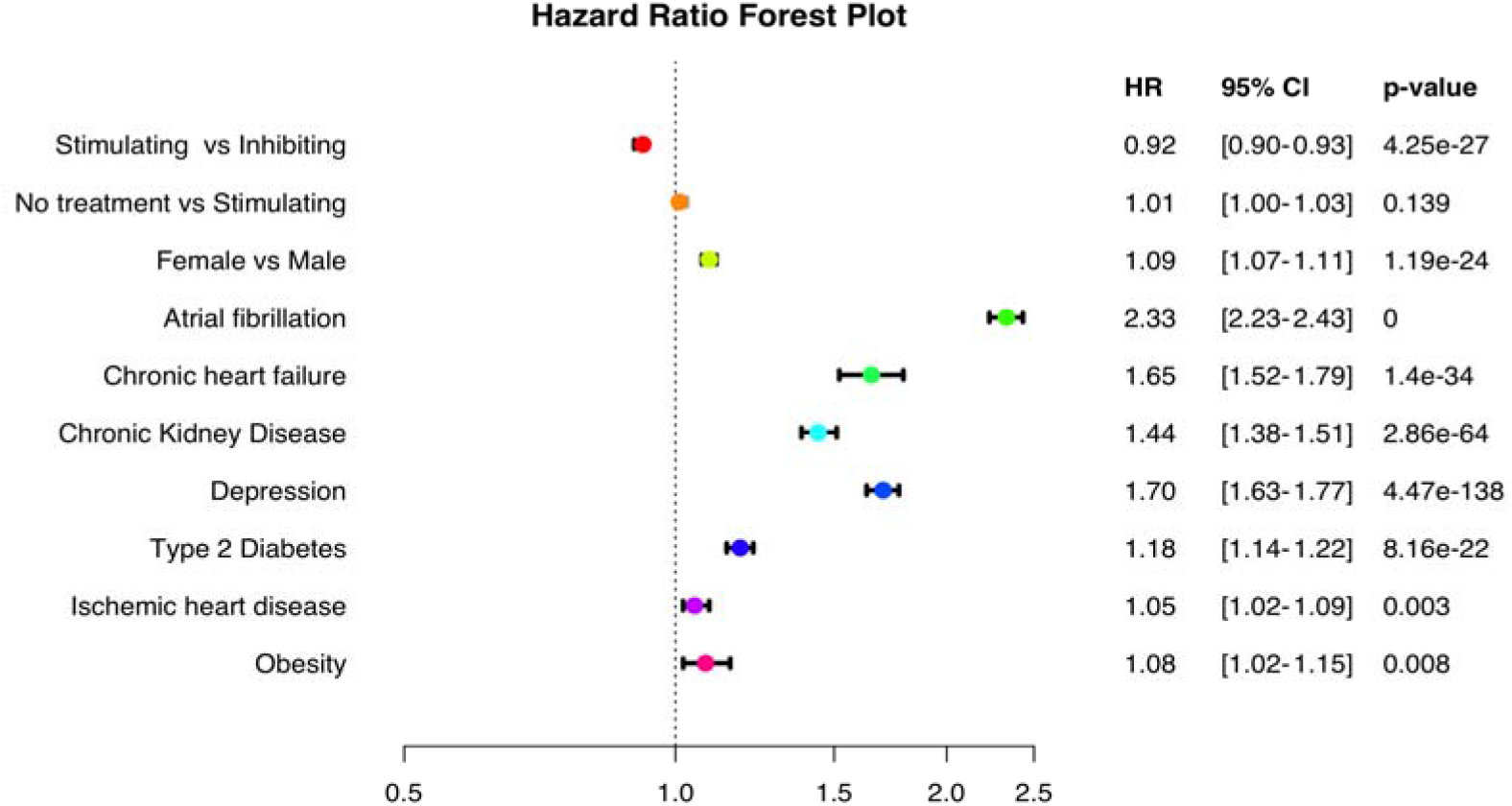
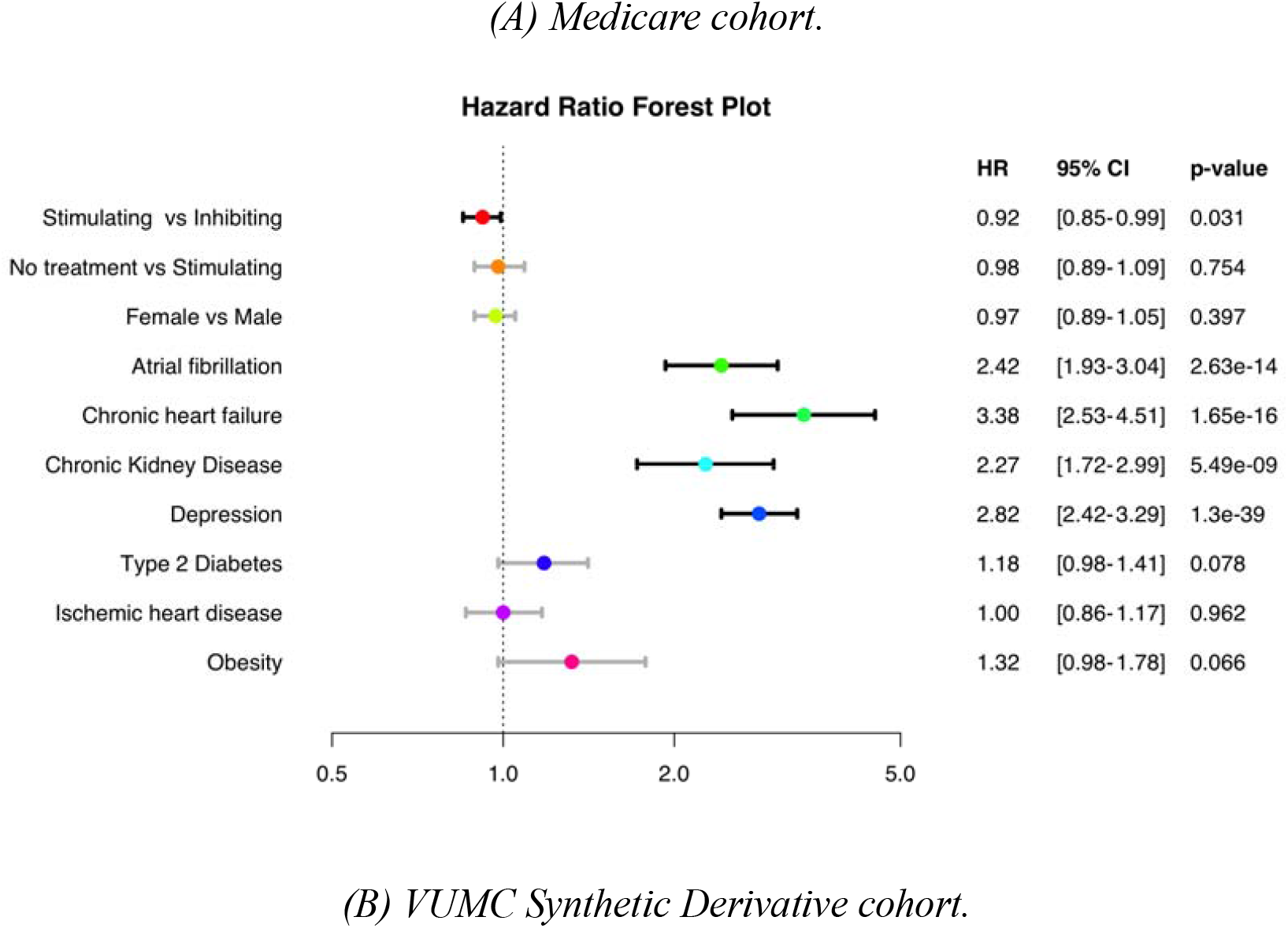
Multivariable Cox proportional-hazards forest plots of adjusted hazard ratios for ADRD by exposure and covariate, in the Medicare (A) and VUMC SD (B) cohorts. Reference for the primary contrast is the inhibiting group; the No-treatment row uses the stimulating group as the reference.

### Replication in VUMC Synthetic Derivative

In the independent VUMC SD replication cohort, predominant use of stimulating antihypertensive medications was again associated with a modestly lower ADRD hazard compared with predominant inhibiting use (adjusted HR, 0.92; 95% CI, 0.85-0.99; *P* = .03), with confidence-interval bounds excluding the null. Nonusers did not differ significantly from stimulating users (HR, 0.98; 95% CI, 0.89-1.09; *P* = .75).

Covariate associations were directionally consistent with established dementia risk factors and with the Medicare cohort, although point estimates were larger, consistent with the smaller, academic-medical-center sample and case mix (Figure 2B). Atrial fibrillation (HR, 2.42; 95% CI, 1.93-3.04), chronic heart failure (HR, 3.38; 95% CI, 2.53-4.51), chronic kidney disease (HR, 2.27; 95% CI, 1.72-2.99), and depression (HR, 2.82; 95% CI, 2.42-3.29) were each strongly associated with ADRD (all *P* < .001). Sex (HR, 0.97; 95% CI, 0.89-1.05) and ischemic heart disease (HR, 1.00; 95% CI, 0.86-1.17) were not significantly associated with ADRD; type 2 diabetes (HR, 1.18; 95% CI, 0.98-1.41) and obesity (HR, 1.32; 95% CI, 0.98-1.78) had elevated point estimates that did not reach statistical significance.

## Discussion

In two structurally independent cohorts of older adults with hypertension, a national Medicare claims cohort drawn from the MarketScan Medicare Database (2015–2023) and an EHR-based academic medical center cohort (VUMC SD), the predominant use of antihypertensive medications that stimulate angiotensin II type 2 and type 4 receptor signaling was associated with a modestly lower risk of incident ADRD compared with predominantly inhibiting agents. Adjusted hazard ratios were directionally and quantitatively concordant across data sources (HR ≈ 0.92 in both), despite differences in patient populations, geographic coverage, and ascertainment mechanisms (claims-based vs EHR-based). This concordance constitutes external replication of the receptor-pathway finding originally reported in a single Medicare cohort by Marcum and colleagues,^26^ and addresses a key limitation of single-database pharmacoepidemiologic studies that estimates may reflect dataset-specific coding, formulary, or case-mix artifacts. The robustness of expected covariate associations, particularly the strong relationships of atrial fibrillation, heart failure, chronic kidney disease, and depression with ADRD, further supports the internal validity of the analytic approach in both data sources.

Our findings are consistent with a growing body of pharmacoepidemiologic evidence implicating differential RAS modulation in cognitive outcomes. Earlier population-based cohorts reported lower dementia incidence among ARB users compared with users of other antihypertensives,^24^ and several subsequent observational studies and meta-analyses have found that RAS-active agents-particularly ARBs-are associated with reduced risk of dementia or cognitive decline,^9,10,11,15^ although class-comparison estimates have varied across cohorts and exposure definitions. The framework adopted here may better capture differential effects on neurovascular biology than conventional drug-class comparisons, because medications grouped within a single conventional drug class can differ in their net effect on AT_2_R/AT_4_R signaling and on cerebral perfusion (eg, dihydropyridine vs nondihydropyridine calcium channel blockers), whereas the receptor-pathway grouping aligns directly with the proposed neurovascular mechanism.^22, 23, 25, 34, 35^

The biological plausibility of these associations is supported by preclinical and translational evidence on cerebral RAS function. AT_1_R activation drives endothelial dysfunction, oxidative stress, and neuroinflammation, whereas AT_2_R/AT_4_R signaling promotes vasodilation, anti-inflammatory effects, neuronal survival, and cerebral perfusion.^16, 18^ By selectively blocking AT_1_R while leaving circulating angiotensin II available to act on AT_2_R and AT_4_R, ARBs may shift net RAS signaling toward neuroprotective pathways. Dihydropyridine calcium channel blockers and thiazide diuretics, which do not directly engage RAS, similarly preserve type 2 and type 4 receptor activity. ACEIs reduce angiotensin II availability across all receptor types and additionally inhibit ACE-mediated amyloid-β degradation, which may attenuate any net cognitive benefit of blood pressure reduction.^16, 20, 36^ Differences in blood-brain barrier penetration among RAS-active drugs may further contribute to heterogeneity across studies.^21^

The principal advance of this study over the prior single-source analysis is the use of two independent healthcare data environments, i.e., a national Medicare fee-for-service claims cohort and an academic-medical-center EHR (VUMC SD) (Medicare claims sourced from the Merative MarketScan Medicare Database), analyzed in parallel with harmonized exposure, covariate, and outcome definitions. In addition, the cohorts in this study are substantially larger (approximately 20-fold larger, given that the prior analysis used a 5% random Medicare sample) than those in the prior study.^26^ Because claims and EHR systems capture prescribing, comorbidities, and diagnoses through structurally different mechanisms (administrative billing vs clinician documentation), concordant estimates across the two sources provide external validation that is difficult to achieve from a single cohort and reduces concern that the association is an artifact of any one dataset’s coding conventions, formulary patterns, or population case mix. The active-comparator new-user design with a 365-day blanking period reduces confounding by indication by restricting the comparison to participants who share the same primary indication for treatment (hypertension), so that the two exposure groups differ principally in the receptor-pathway class of antihypertensive used rather than in underlying disease; propensity-score matching produced excellent baseline balance within each data source, reducing the influence of measured confounders; and a long follow-up window in both cohorts (approximately 7 years) increases statistical power to detect modest associations with incident ADRD. The receptor-pathway exposure framework links the analytic contrast to a specific biological mechanism (AT_1_R blockade with preserved AT_2_R/AT_4_R activity) rather than to administrative drug-class labels.

Several limitations warrant consideration. First, residual confounding by indication or by unmeasured factors (e.g., frailty, prescriber preference, prodromal cognitive symptoms not captured by claims) cannot be excluded; observational pharmacoepidemiologic effect sizes of this magnitude are within the range potentially explained by unmeasured confounding. Second, exposure was ascertained from prescription fills rather than verified intake, so nonadherence is likely nondifferential misclassification. Third, ADRD is incompletely captured in administrative data; we used coding consistent with the CMS Chronic Conditions Data Warehouse definition to mitigate misclassification but cannot exclude differential ascertainment between cohorts. Fourth, the VUMC SD represents a single academic medical center and may not generalize to other care settings. Finally, while estimates were directionally and quantitatively concordant across cohorts, observational data alone cannot establish causality. Despite the limitations, the validation of the results across two structurally distinct EHR systems in this study mitigates these challenges.

Hypertension affects approximately 1.28 billion adults globally and is among the most prevalent treatable midlife risk factors for dementia.^9, 37^ Even small relative reductions in dementia risk associated with class selection could translate into meaningful population-level effects, particularly given the growing prevalence of dementia in the aging population.^38^ If confirmed in randomized comparative-effectiveness trials, these findings could inform pragmatic prescribing decisions for older adults with multiple guideline-equivalent antihypertensive options.^39^ Future research should evaluate effect modification by age, sex, race, and ethnicity, and APOE genotype;^40^ pursue head-to-head randomized trials with cognitive endpoints, complemented by multidomain lifestyle intervention approaches;^41^ and integrate neuroimaging and fluid biomarker outcomes to clarify mechanisms of differential RAS modulation in neurodegeneration.

## Conclusion

In two large, structurally independent cohorts of older adults with hypertension, predominant use of antihypertensive medications that stimulate angiotensin II type 2 and type 4 receptor signaling was associated with a modestly lower risk of incident Alzheimer disease and related dementias compared with predominantly inhibiting agents. Concordant estimates across data sources (HR ≈ 0.92 in both) externally replicate prior single-database evidence and strengthen its validity by demonstrating consistency across structurally distinct healthcare data environments. Although causal inference cannot be drawn from observational data, the consistency of estimates supports targeted randomized comparative-effectiveness trials of receptor-pathway–informed antihypertensive selection for cognitive outcomes.

## Data Attribution

Certain data used in this study were supplied by Merative as part of one or more MarketScan Research Databases. Any analysis, interpretation, or conclusion based on these data is solely that of the authors and not Merative.

## Acknowledgement

The study was supported by NIH funding R01AG069900.

## Article Information

### Accepted for Publication

____________________.

### Author Contributions

B.L., A.T., Z.W., W.Q.W., and X.Z. Conceived and supervised the study. A.T. collected and performed the EHR analysis. B.L and A.T. wrote the manuscript. All authors participated in the interpretation of the results and read and/or edited the final manuscript. All authors approved the final version of the manuscript and consented to publication.

### Data Sharing Statement

The MarketScan Medicare Database is proprietary to Merative and cannot be shared publicly by the authors; researchers may obtain access through a data-use agreement directly with Merative. VUMC Synthetic Derivative data are available to qualified investigators through the Vanderbilt Institute for Clinical and Translational Research pursuant to institutional data-use policies. Aggregated deidentified analytic datasets, code lists, and analytic code supporting this analysis are available from the corresponding author upon reasonable request.

### Supplement 1

eTable 1. ICD-9 and ICD-10 hypertension diagnosis code list used for cohort identification. eTable 2. Full list of qualifying antihypertensive medications by receptor-pathway class.

Trademarks. Merative and MarketScan are trademarks of Merative Corporation in the United States, other countries, or both.

