## Supplement 1 for "Antihypertensive Medication Class and Incident Alzheimer’s Disease and Related Dementias: External Replication Across Two Independent Healthcare Data Sources-the Vanderbilt EHR Synthetic Derivative and US Medicare Claims": Supplement_1.docx

**eTable 1.** *ICD-9* and *ICD-10* Diagnosis Codes

| **Disease** | **ICD-9 Codes** | **ICD-10 Codes** |
| --- | --- | --- |
| **Type 2 Diabetes** | 250, 250.02, 250.1, 250.12, 250.2, 250.22, 250.3,  250.32, 250.4, 250.42, 250.5, 250.52, 250.6,  250.62, 250.7, 250.72, 250.8, 250.82, 250.9, 250.92 | E11.0, E11.1, E11.2, E11.3, E11.4, E11.5,  E11.6, E11.7, E11.8, E11.9 |
| **Alzheimer's Disease / Dementia** | 331,331.11,331.19,331.2,331.7,290,290.1,  290.11,290.12,290.13,290.2,290.21,290.3,  290.4,290.41,290.42,290.43,294,294.1,294.11,  294.2,294.21,294.8,797 | F01.50,F01.51,F02.80,F02.81,F03.90,  F03.91, F04,F05,F06.1,F06.8,G13.8,G30.0,  G30.1,G30.8,G30.9,G31.01,G31.09,  G31.1,G31.2, G94,R41.81,R54 |
| **Hypertension** | 401,401.1,401.9,402,402.01,402.1,402.11,402.9,  402.91,403,403.01,403.1,403.11,403.9,403.91,  404,404.01,404.02,404.03,404.1,404.11,404.12,  404.13,404.9,404.91,404.92,404.93,405.01,  405.09,405.11,405.19,405.91,405.99 | I10,I11.0,I11.9,I12.0,I12.9,I13.0,I13.10,  I13.11,I13.2,I15.0,I15.1,I15.2,I15.8,I15.9 |
| **Obesity** | 278,278.01,278.02,278.03 | E66.0,E66.01,E66.09,E66.1,E66.2,E66.3,E66.8,E66.9 |
| **Chronic Kidney Disease** | 585.1,585.2,585.3,585.4,585.5,585.6,585.9 | N18.1,N18.2,N18.3,N18.4,N18.5,N18.6,N18.9 |
| **Depression** | 296.2,296.21,296.22,296.23,296.24,296.25,296.26,  296.,296.31,296.32,296.33,296.34,296.35,296.36,  300.4,311 | F32.0,F32.1,F32.2,F32.3,F32.4,F32.5,F32.8,  F32.9,F33.0,F33.1,F33.2,F33.3,F33.4,F33.8,  F33.9,F34.1 |
| **Ischemic Heart Disease** | 410,410.1,410.2,410.3,410.4,410.5,410.6,410.7,  410.8,410.9,411,411.1,411.8,411.81,411.89,412,  413,413.1,413.9,414,414.01,414.02,414.03,414.04,  414.05,414.06,414.07,414.8,414.9 | I20.0,I20.1,I20.8,I20.9,I21.0,I21.1,I21.2,  I21.3,I21.4,I21.9,I22.0,I22.1,I22.2,I22.8,  I22.9,I23.0,I23.1,I23.2,I23.3,I23.4,I23.5,  I23.6,I23.8,I24.0,I24.1,I24.8,I24.9,I25.0,  I25.10,I25.11,I25.110,I25.111,I25.118,  I25.119,I25.2,I25.3,I25.4,I25.5,I25.6,  I25.7,I25.8,I25.9 |
| **Atrial Fibrillation** | 427.31 | I48.0,I48.1,I48.2,I48.3,I48.4,I48.9,I489.1 |
| **Congestive Heart Failure** | 428,428.1,428.2,428.21,428.22,428.23,428.3,  428.31,428.32,428.33,428.4,428.41,428.42,  428.43 | I50.1,I50.20,I50.21,I50.22,I50.30,I50.31,  I50.32,I50.40,I50.41,I50.42 |

**eTable 2.** Antihypertensive Medications by Category

| **Angiotensin II type 2 and 4 stimulating** | **Angiotensin II type 2 and 4 inhibiting** |
| --- | --- |
| Azilsartan; Candesartan; Eprosartan; Losartan; Olmesartan; Telmisartan; Valsartan; Irbesartan; Amlodipine; Felodipine; Nicardipine; Nifedipine; Nisoldipine; Isradipine; Nimodipine; Chlorothiazide; Hydrochlorothiazide; Indapamide; Bendroflumethiazide; Chlorthalidone; Hydroflumethiazide; Methyclothiazide; Polythiazide; Trichlormethiazide | Benazepril; Captopril; Enalapril; Fosinopril; Lisinopril; Moexipril; Perindopril; Quinapril; Ramipril; Trandolapril; Acebutolol; Atenolol; Betaxolol; Bisoprolol; Carvedilol; Labetalol; Metoprolol; Metoprolol tartrate; Nadolol; Nebivolol; Pindolol; Propranolol; Carteolol; Penbutolol; Sotalol; Timolol; Diltiazem; Verapamil; Bepridil; Mibefradil |
